## Supplementary material for "Conceptual framework of episodic disability in the context of Long COVID: Findings from a community-engaged international qualitative study": S1-Interview Guide

**Supplemental File 1 - Interview Guide**
**Episodic Disability and Long COVID Study**

**Preamble**: Thank you for agreeing to participate in this study. I am meeting with you to try to get a better understanding of the health-related challenges (otherwise known as disability) you might experience living with Long COVID. I would also like to understand the extent to which these challenges faced by someone living with Long COVID are fluctuating (or episodic) in nature.

Disability can be defined in several ways. It may include difficulties with your physical, mental, or emotional health, or difficulties with your cognition or thinking. Disability may also encompass difficulties with day-to-day activities, challenges to social inclusion, and uncertainty or worrying about the future that you may experience living with Long COVID. We want to learn more about what these health-related challenges are, and how they might fluctuate – on a daily basis as well as over weeks or months. I am also interested in how predictable your health challenges are, whether you know the cause of your episodes of health challenges and what (if anything) triggers them or helps them improve. Finally, I am interested in hearing about the strategies or services you use to deal with the periodic ups and downs of disability living with Long COVID.

Your experiences and perspectives are important. Results from these interviews will be used to serve as a foundation for understanding and measuring episodic disability in Long COVID. Lastly, please remember to let me know if you'd like to take a break at any point during this interview.

Do you have any questions before we begin?

A) **Dimensions and Experiences of Disability**

**Can you describe any health-related challenges that you currently experience or have experienced living with Long COVID?**

*Probes*: Any physical health challenges?
Any mental health or emotional health challenges?
Any memory or cognitive health challenges?
Any difficulties carrying out day-to-day activities?
Any challenges to social inclusion (work, employment, income, relationships (friendships, intimate relationships, partnerships), caring for others, caring for children)?
Any challenges that pertain to worrying about your future health or uncertainty living with Long COVID?
Any other health challenges?

B) **Episodic Nature of Disability**

**Can you describe how your challenges might change over time living with Long COVID?**

Are there some health challenges that come and go, and others that are more constant?

*Probes*: Any fluctuations in physical health?

Fluctuations in fatigue, post-exertional symptom exacerbation/malaise, muscle/joint/chest/abdominal pain, sensation, lightheadedness/dizziness, skin rashes, or sense of taste and smell?

Any fluctuations in mental health or emotional health?

Fluctuations in mood?

Any fluctuations in memory or cognitive health?

Fluctuations in ability to think or concentrate?

Any fluctuations in ability to carry out day-to-day activities?
Any fluctuations in social inclusion (work, employment, income, relationships)?
Any fluctuations in your worrying about your future health or uncertainty living with Long COVID?
Any other fluctuations?

Can you walk through what might be a typical episode of health / disability?

Are the fluctuations in health predictable / unpredictable? Can you explain?

How severe are the 'ups and downs' of disability that you experience?

How frequently do you experience your 'ups and downs' or changes in disability?

How long (duration) might you experience episodes of disability?

What is the impact of these episodes (severity, frequency, duration) on your overall well-being?

*Probes*: Employment
Access to services
Financial security
Relationships
Caretaking responsibilities

What about the episodes of disability do you find most challenging?

Illustrating Episodic Experiences: Can you draw what your level of health living with Long COVID has been like? Can you draw what a typical day is like living with Long COVID in relation to episodic disability? [*participant draws on White Board – Annotation in Zoom*].

### C) **Uncertainty and Episodic Disability**

**Do you experience uncertainty living with Long COVID? If yes, please explain.**

Probes: Worried about when next episode might arise?
Worried about the severity of the episode?
Worried about what the consequences of the episode might be for yourself?
Worried about what the consequences of the episode might be for others?
Worried about the cause of Long COVID in general and whether it will ever get better?

Do you have some days where you feel more uncertain than others or is the uncertainty there all the time?

Do you have any concerns about the future living with Long COVID?

### D) Triggers of Disability

**What sorts of things might trigger an episode of illness or disability for you?**

*Probes*: Exercise
Stressful event
Screen time
Invalidation or disbelief of symptoms during health care access event
Menstruation
Mental strain

Do you know why you experience these ongoing symptoms and challenges of Long COVID while others may not?

Do you feel it is important to you to know the cause or source of your disability?  If yes, does not knowing increase your uncertainty living with Long COVID?

### E) Extrinsic and Intrinsic Contextual Factors that Influence Disability

**Is there anything in the environment that influences your health related challenges (or disability) that you experience living with Long COVID?   Anything that makes the health challenges you experience better or worse?**

*Probes*: Health / rehabilitation services / providers
Community support networks
Stigma
Support from family / friends
Financial / employment policies / benefits
Family caring roles and responsibilities, financial

**Is there anything personal/specific to you that you think might influence your health-related challenges (or disability) that you experience living with Long COVID?   Anything that makes the health challenges you experience better or worse?**

*Probes*: Other health conditions you might be living with in addition to Long COVID
Age
Gender
Sex
Race/ethnicity
Employment status
Length of time living with Long COVID

Severity of initial COVID-19 infection
Vaccination status when you were diagnosed with COVID-19
Treatment you did or did not receive during initial illness

F) **Living Strategies to Deal with Episodic Disability**

**Have you ever accessed any health (including mental health), rehabilitation (e.g. occupational therapy, physical therapy) or social services to help you with your health challenges living with Long COVID?**

If yes, can you tell me about your experience accessing these services?

Why did you access? How did you access? Where did you access? Did you specifically seek out a Long COVID clinic? When did you access? Who did you see? What kinds of treatment or services did they provide?

Did you find these services were helpful in reducing or preventing the number of health challenges that you might experience?

Did you feel that your challenges were adequately acknowledged and addressed by the services you accessed?

Did you experience any challenges to accessing services?

Did you adopt any other living or coping strategies to help you deal with the health challenges and ups and downs of disability living with Long COVID?

If so, what were they?

*Probes*: Maintaining control over health as able (e.g. pacing, healthy lifestyle, nutrition, sleep, prioritizing)
Attitudes and Beliefs
Trying to block out living with Long COVID altogether?
Seeking out social interaction / reaching out for social support from others

Did you find any of the strategies were helpful in reducing or preventing disability?

**Summary**

Do you have anything else you wish to say about the health-related challenges / episodes of disability you experience living with Long COVID?

Thank you very much for participating in this interview today. Your responses will help to develop a future tool that we can use to measure Episodic Disability among adults living with Long COVID. We hope this will help us better understand the presence, severity and episodic nature of disability experienced by adults living with Long COVID.

Next Steps: In the next phase of this study, we will be administering a questionnaire on Episodic Disability via email to adults living with Long COVID. The questionnaire will be based on the findings from these interviews. The next phase of the study involves completing a series of questionnaires (including an Episodic

Disability Questionnaire) via a survey link that will be emailed to you, followed by completing the EDQ only 1 week later (also by a survey link in an email).

Would you be willing to be contacted about participating in the next phase of the study?
