## Supplementary material for "Conceptual framework of episodic disability in the context of Long COVID: Findings from a community-engaged international qualitative study": S2-Dimensions of Disability - Supportive Quotes

**Supplemental File 2:** Dimensions of Disability in the Episodic Disability Framework in the Context of Long COVID - Supportive Quotes (n=40 participants)

| Dimension of Disability | Health challenges that comprise the dimensions | Supportive Quotes |
|---|---|---|
| **Cognitive Symptoms and Impairments** | Post-exertional symptom exacerbation | Now I crashed and I remained in that post exertional symptom exacerbation state for a number of days. So then it just became about survival. It just became about literally lying in bed and feeding yourself. And then you have to work really hard to get yourself out of it. You've got to do your breathing and your meditation and your tens machine and you know make sure that you get sleep. But sometimes it just happens for no reason and that is very frustrating. – P31, UK<br><br>I did engage with some physiotherapy offered by [the Long COVID clinic] but that was… also I kind of stopped it because I was having to one go into the centre. So I was having really bad pain afterwards and then it meant I couldn't come home and cook dinner for example. – P1, UK<br><br>So fatigue for probably all of 2020 was I would… if I overdid it, I would find that two to three days later I would kind of go through like a crash. So I would feel the need to sleep more. Waking up in the morning I'd have lower energy. My mood would be off just because I didn't feel well rested. Even though if I slept like nine to ten hours I just didn't feel like well rested. Like I didn't feel recharged. – P27, Canada |
| | Cognitive Endurance | If I want to send an email, something I would have done at work without thinking about it, now it could take the whole day of energy to build up to be able to do it. It's not the physical thing. It's an exhaustion from I suppose the cognitive side of it. It drains me to do it to try to figure out what to write, how to write it, how to send it, have I sent it to the right email or not, second guessing myself because I make so many mistakes. So those things have worsened since the second bout of COVID. – P36, Ireland<br><br>But again, it doesn't take a whole lot to flare up some of the cognitive symptoms as well. – P2, US |
| | Executive Function | The brain fog, like I said to the consultant there will come a time when I will sit down and I will describe what it is because it's not a fog. It's like being completely lost. It's like being slightly removed from who you are. You can see everything you need but it's out of reach. You can't get to it. It's like… so I suppose it is like being lost at sea in a fog literally. But it's profoundly distressing and profoundly confusing and it is like having dementia. It is like… because you really feel like you're not coping and it's just not even to be able to |

**Supplemental File 2:** Dimensions of Disability in the Episodic Disability Framework in the Context of Long COVID - Supportive Quotes (n=40 participants)

| Dimension of Disability | Health challenges that comprise the dimensions | Supportive Quotes |
|---|---|---|
| | | write an email or not even to be able to fill out a form for yourself, to ask for help for yourself, not to be able to manage your thought process well enough to be able to do a thing like that. That for the likes of me who used to be a school Principal and so busy and multitasking, now I have to like… to cook for myself or to make bread because I'm on this really strict diet through the consultant, even to be able to make the bread, like my sister is coming, stir the ingredients for me or chop up the things I need. – P12, Ireland<br><br>I'm not sure what it is about the supermarket. Is it the trying to figure out what I need off my list, what to put in the trolley… so I can't believe I even find this effort. – P8, Ireland |
| | Word finding, and communication (written and oral) | By the time I could sit comfortably and write, I was… I basically couldn't… I would be thinking what I'm going to write and then I would go to write it down and it wasn't what was coming out. It was like I was making… and I used to be like perfect spelling across the board. Like every single time in my class, always number one. This would be like jumbled letters and words and I would write like two letters and I'd have to stop. I'd be like okay it's not flowing, like I can't get it out. Then weird things like I would be… I would want to make a post on Facebook or Instagram or something and I'd write it out. I'd read it like a million times because I was paranoid like I don't you know… because of like how bad cognitively it was, I was worried that I was making mistakes. I'd read it like 10 times and then I'd post it and then suddenly I'd look back at it half an hour later and read it and be like this makes no sense. Like even with me checking it, it was so difficult to get what was in my head on a page. – P18, Canada<br><br>…and there were cognitive impacts. Like you started off with just word finding difficulties. – P19, UK |
| | Concentration | So before getting COVID I was a full-time teacher. I was a history teacher. Now I'm forgetting things constantly. I'm relying constantly on other people to help me remember things. My concentration is appalling. Like after reading say five pages of a book, I can't read anymore because I just feel kind of overwhelmed by that. So yeah, that's been one of the most difficult things to kind of deal with. – P32, UK<br><br>Probably the first seven months, there was a fair amount of brain fog, a fair amount of difficulty concentrating. I want to say that there was some memory issue for a little while. I think the memory issues have improved and the brain fog has improved significantly. – P2, US |

| Dimension of Disability | Health challenges that comprise the dimensions | Supportive Quotes |
|---|---|---|
| | | Sometimes I set a reminder when I'm working to stop after 30 minutes and do nothing and maybe just listen to the radio for 15 minutes. You know I don't… any time I go more than 30 minutes steady focus things get worse, you know depreciate more rapidly. So it's 30 minutes, okay stop, break and then pick it up. – P15, US |
| | Memory (short-term; long-term; working) | I suppose the other part would be the cognitive decline. That one is very frightening. I would… it's not consistent. It's kind of random word loss. My short term memory is very poor. So even though… like say my story and my journey of Long COVID is very much with me and current you know because I'm in it, I still have to have all my notes next to me because I could go blank in the midst of it and I might not… I don't know. I never know what way that would be. – P36, Ireland<br><br>In my most recent relapse the one I'm in kind of like right now, that was when I really saw some of the cognitive issues. So recall, like short term memory, losing my words, struggling to like track… yeah, I just like follow conversations, that kind of thing. I think that yeah… and like feeling brain fog. The brain fog was really prevalent. – P35, US |
| | Information processing; Speed of Processing | I have to keep re-reading every reading. So I've cut out every other thing. I used to love to read and I'd read two or three books a month. I'm now at the same book for two years and I haven't got past the first few chapters because I can't retain any of it. – P36, Ireland<br><br>On any given day I can start… you know I can do some work and if I don't pace it correctly, by 2pm or 3pm I've got a really bad headache and really bad nausea and I just lie and listen to some music or something. I have to shut myself down after a couple of hours of what I would call engaged work. I think of if I'm doing analytic work involving pivot tables or setting up PowerPoint slides, thinking about how to present information, that can… I have about 90… if I can space out a couple of you know sessions, I can usually get about 90 minutes of quality work out of myself. – P15, US<br><br>…like one small change is being more honest with myself about how my symptoms are in control or not. So previously I would be like well I'm doing more physically and I want to keep doing more, therefore you know we're going to run down. But yeah, just becoming more honest with myself and also more aware I guess of my gains. So like if previously I really struggled to make inferences on a 30 minute call and now I'm able to make |

| Dimension of Disability | Health challenges that comprise the dimensions | Supportive Quotes |
|---|---|---|
| | | inferences but I still can't go beyond 30 minutes, that's still a victory. Just being more cognizant of that has also impacted my thinking. – P17, US |
| | Multi-tasking | I mean I was an art director and a producer and a director. Like I was in costume design. I was doing all these things and I was working you know tons of hours, running teams of people and lifting stuff, moving stuff. I mean I was travelling the world all over the place, film festivals. I was more full blast than anybody that I knew in terms of doing things and being active and getting stuff done and multitasking. It's gone from like 1000% to like 5%. So I'm now kind of… I am kind of going through this like grieving process of my like previous self. – P18, Canada<br><br>I think the fatigue and the… so even besides word finding difficulties, you know just the not being able to follow a straight line. I can't… so like today I got up. I felt really energetic. I knew we needed toilet rolls upstairs but I didn't remember that again until after lunch. So everything I… every job that I go to do, I get distracted by something else and I do something else and then the jobs don't get done you know. So hence you know I put banana bread in the oven but then I completely forgot about it. So it's just like that with everything you know, all of the jobs. So sometimes they get done and sometimes they don't. There was washing in the machine since last night that I had forgotten about that I stumbled across about 11 am. So I put it on to dry. So everything's a bit higgledy piggledy really. – P8, Ireland |
| **Mental and Emotional Symptoms and Impairments** | Grief, devastation, loss (function, health, relationships, employment, career, finances) | Before you called I was listening to a daily episode of the New York Times and it was saying you know we're doing an episode on people who've lost someone from COVID and if you had anything that you want to say about someone you lost from COVID, call. The first thing I thought was I want to do an episode like that with Long COVID patients who've lost their lives because we've all lost our lives and no one …. understands that all our lives have been destroyed (crying). Like they've been completely destroyed and no one… like people are walking around like it's nothing." – P18, Canada<br><br>I think… I think that… I think about how it is unpredictable in terms of the amount, how it affects the entire body and how it can cross between… and it changes. So whether that's cognitively where for a period of thankfully it was a month or two my perception of the world changed like as if someone had given me a drug that much like. So when that happens, that was very scary because I felt a loss of sense of who I was and I felt like I |

| Dimension of Disability | Health challenges that comprise the dimensions | Supportive Quotes |
|---|---|---|
| | | needed somebody to advocate for me and there was no one. I had to do it for myself. I felt quite vulnerable then. – P3, UK<br><br>Realizing that I couldn't work …towards the end of when I was in my job and kind just realizing I really couldn't do it anymore. That was really devastating because I liked my job and I worked really hard to get that job and I had a lot of, still have a lot of student loans, you know, just like the financial implications of that. My partner and I really prioritized my career as like the income we were mostly relying on. So that was just practically speaking very stressful but then also you know it was emotionally sad. Like I put a lot into my career. – P26, US |
| | Anxiety or depression | …feeling very low, not feeling myself and obviously feeling a lot of anxiety about everything like what am I doing, why am I even working, how could they give me… you know like how could they let me work when I can't even understand what people are saying in a meeting. You know these sorts of like anxieties. – P21, UK<br><br>I've never never suffered with anxiety before in my life. I've not been a nervous person. I've not… I was the one that's always been bubbly, out there, I would try anything, adrenaline junky, that type of thing. I used to think… I've got family members that suffer with anxiety and they used to always ring me up because I was the healthcare professional and I'd be like 'you're overthinking, you're overthinking.' But when you actually deal with it yourself it's such a different… I totally kind of resonate with my family members now. I apologized. I said I really apologize for going 'yeah, you're alright, you'll be fine'. It takes its toll on you. So yeah. – P4, UK |
| | Fear | I think you get a fear in your gut that it's just the way it's going to be the whole time. Some of our group there, 17 and 18 months and I'm still on 11 months and I'm just… I have that fear like. As my doctor says, he keeps reassuring me that maybe after 18 months I might start to… but we have to keep hope. – P10, Ireland<br><br>I think in a weird way like accepting the uncertainty is a freeing feeling because you kind of become less invested in like the outcome. But the actual feeling of the uncertainty is terrifying. – P18, Canada<br><br>Honestly, even if I could… even if suddenly and miraculously I were approved for ODSP like tomorrow, which is not going to happen, I don't know how anybody lives on it. The |

| Dimension of Disability | Health challenges that comprise the dimensions | Supportive Quotes |
|---|---|---|
| | | idea with ODSP is that you get $1,169 [CAD] a month for rent, food, clothes and whatever you need. There's actually nowhere to live anywhere in Canada with that kind of money. And then they're like 'you can get a part time job' and I'm incapable of getting any kind of meaningful or you know… I can probably work an hour and a half every three days if I really tried hard and that job doesn't exist. So in that case you know I could get ODSP. I would still eventually become homeless and then they take away your shelter portion. So then … you have $600 a month to live on the street if you can imagine. So I don't have any financial uncertainty. What I have is a terrible fear of like homelessness. – P22, Canada |
| | Anger, mood swings, irritability | You have no control over your emotions and you just… it's like you're manic depressive and you just go crazy, crazy. It's very common in long haulers and I heard kids are also getting it as well. – P13, Canada<br><br>I think it would look pretty similar or you would see patterns where if I'm thinking about my mood would kind of be at [not audible 63.10] really low points. I would also say that then if I were to graph my… like knowing that like certain periods of emotion and anxiety which for me is kind of takes a lot of energy precipitates one of these, it would maybe even be a little higher than the energy bar and then go down lower. But I think it would follow that same kind of manner. – P24, US |
| | Hopelessness; suicide ideation | I'm not exaggerating, I was suicidal. That's how bad I was. I never knew anything. I didn't know until my depression. I mean I had down days like everybody else but this was so traumatic and I was so sick. Yeah, I got very very depressed and I was suicidal. The only thing that kept me going was that there are a few grandchildren but my 10 year old, the eldest grandchild is about 10. We lost two family members to COVID in March 2020 within 10 days of each other and we all got sick. She got very anxious about this. Only I knew she wouldn't cope if I had done something. That's the only… myself and me husband are married 40 years. We still get along great and I have two lovely kids. They're married and they've got kids and I figured they'd get over it. But the 10 year old was so bad. I thought I can't do that to that child, it'll ruin her life. So I went to my GP and he put me on antidepressants. I started psychotherapy on zoom as well and I found a fantastic woman. So the two of them saved me. I started to feel a bit better in myself. I was still full of despair but I wasn't in that dark place. – P7, Ireland |

| Dimension of Disability | Health challenges that comprise the dimensions | Supportive Quotes |
|---|---|---|
| | | I don't like saying it but I've been suicidal just kind of like you know what kind of life is it. You know I just sat in the toilet for you know 10 hours a day and I was like what is this. I sat there for like you know 10 weeks you know. No way of life to sit on the toilet, go to bed, sit on the toilet, go to bed and like what kind of life is this you know when you were so active pre COVID and things. I started medication for anxiety which has been increased. – P20, UK |
| | Guilt | I do feel guilty for my kids. You know sometimes they want a piece of me and they're teenagers. So they don't often come. It's not like they're little ones who are crawling all over you. It feels like it's a really hard balance between meeting their needs and saying actually I've got to look after myself now and I can't do anymore. As they're teenagers their issues are bigger and more complex. – P19, UK<br><br>So there's a big nobody knows and I'm kind of like… I almost feel guilty when I call the doctor saying like oh this is going on now or if I've got some new symptoms or most of my new symptoms I don't even tell my doctor because I'm just like they're bearable and I don't want to keep bugging which he does always say you're not bugging, just call. – P30, UK |
| | Loneliness | Yeah, being alone is not ideal for people. You know we're not kind of built for… you know it's nice to have quiet time. But you know there… you know there are very few real hermits in the world. Most people need to be around at least a couple, one or two other people. So being… you know being alone I think has been not the best for my mental health. … and not the best for my physical health because there are times when I shouldn't be doing things and I have to do them because there aren't other people to help. Yeah, being isolated…. being left alone with your own thoughts for that long is not good for one person. It's not good for your mental health or even your physical health is affected. Emotionally it's anxiety-making, it's depressing. – P22, Canada<br><br>I'm just glad I even can talk to you and you're my human contact for today. I don't have no other contacts for the rest of the day. That's why I just keep talking to you. It's like there's nobody else. I have nobody else for the rest of the day. – P39, Canada |

**Supplemental File 2:** Dimensions of Disability in the Episodic Disability Framework in the Context of Long COVID - Supportive Quotes (n=40 participants)

| Dimension of Disability | Health challenges that comprise the dimensions | Supportive Quotes |
|---|---|---|
| **Difficulties with Day-to-Day Activities** | Mobility (ambulation, stairs), sitting upright | But oh yeah I was telling you when I had the lung infection in January, I slept downstairs for two weeks. I have a bed there. I was not able to go up my stairs. Like there used to be a time when just brushing my teeth set off a tachycardic response, just moving, just getting up off the couch. But now I'm much better, so I actually haven't had an episode in the last two weeks. – P12, Ireland<br><br>So it's like these things that I used to just take for granted like going for walks, going skiing, you know sitting at a desk. You know those aren't… so sitting in a restaurant is not an option. If I go to a restaurant I have to find a place where I can… you know a booth where I can sit sideways and lean so I'm mostly reclined. – P15, US |
| | Bathing, showering, dressing, brushing teeth | I was lying down all the time at home and was barely able to get what I needed to eat. So you know I was showering the bare minimum. Like now I still only have a bath once a week. Like it's bare minimum management of your daily needs you know. – P12, Ireland<br><br>But I had an extremely mild illness, like no fever, no respiratory involvement. I've had worse colds in my life. So I think most of the people in that camp kind of have a similar experience as me where we just get worse when we do stuff which is so hard because having to realize that doing stuff means loading the dishes in the dishwasher, getting dressed, brushing your teeth, you know taking a shower, feeding your cats, like things that you just didn't have to think about before. Now you realize they actually take energy and we don't have energy. – P34, US |
| | Meal preparation, cleaning | I couldn't cook myself meals. I couldn't clean the house. I was bed-bound and couldn't actually do anything. Because sometimes I was either having trouble walking or when I would stand up my heart would you know just be like jumping you know through the roof. So it's changed my life dramatically. – P18, Canada<br><br>My partner was not employed and currently is not employed. So that has been really stressful because it's basically like you know… they have to do everything for me now which they do and I'm… I mean that's actually a bonus or like a positive environmental factor. Like I have live-in help. Like if I can't cook… I don't clean. I don't go grocery shopping. – P26, US |
| | Shopping (e.g. groceries) | Going to the supermarket. I'm not sure what it is about the supermarket. Is it the trying to figure out what I need off my list, what to put in the trolley and then the actual physical… so I can't believe I even find this effort. But the actual putting the stuff into the |

| Dimension of Disability | Health challenges that comprise the dimensions | Supportive Quotes |
|---|---|---|
| | | trolley, getting the stuff off the trolley onto the belt, back into the trolley and into the car and then into the house at home. That's five times that you're taking stuff in and out. I can't believe that's even an issue that I can see that that's a problem. So you know I'm doing a lot of [Brand of Store] online shopping and stuff like that and click and collect. But even the click and collect is hard because I'm still taking stuff in and out of the car and at home. – P8, Ireland<br><br>It's not like I can just like say okay I rested yesterday, so I'll be fine today. That's not the way it is. It's just that I'd have a better chance of being able to have this chat with you. Whereas if I used up a lot of energy yesterday, if I did… like if I went shopping yesterday, you know I might not be as able to do this today. It's just knowing those things. It's kind of you start learning that's the way it is. – P9, Ireland |
| | Going to appointments | Yeah and I suppose just like yeah, living in the country. Now like you have to drive to places. Trying to get to the hospital, I can't drive myself to the hospital anymore because there's no… like there's no services anywhere for long COVID anywhere near where I live. So I have to travel to Dublin for the clinic I'm attending. It's not even a long COVID clinic. It's under infectious diseases. But yeah, I have to get somebody to drive me there. – P35, Ireland<br><br>I know that if I got out twice in two days or do something that's vaguely taxing twice in two days, which is for me yesterday and today because I had a GP appointment and today I'm seeing you, I know that my weekend will be a rubbish weekend. So I can predict that. – P19, UK |
| **Challenges to Social Inclusion** | Recreation; leisure and other social interactions and activities | So it's like these things that I used to just take for granted like going for walks, going skiing, you know sitting at a desk. You know those aren't… so sitting in a restaurant is not an option. If I go to a restaurant I have to find a place where I can… you know a booth where I can sit sideways and lean so I'm mostly reclined. So all those social things just are out. If we have friends over I can engage for half an hour, 45 minutes or maybe a little longer. But I've come to a point where I just say no, I've got to go upstairs, I've got to go lie down. So all of those things basically are just dramatically different. – P15, US<br><br>I was able to get back to activity again pretty quickly or not quickly but comparatively since my last relapses it felt quickly. I have a dog. So I was able to get back into like |

**Supplemental File 2:** Dimensions of Disability in the Episodic Disability Framework in the Context of Long COVID - Supportive Quotes (n=40 participants)

| Dimension of Disability | Health challenges that comprise the dimensions | Supportive Quotes |
|---|---|---|
| | | walking and I'm in a dance class and I was able to do that too. Then I experienced several relapses. – P35, US |
| | Personal relationships | I'm really lucky. My partner has been brilliant. He works from home. His income is such that we can take mine not working. It's not putting us in any difficulties. So I feel you know we are really really privileged from that point of view. All this to say, it's made our relationship stronger. I've had to be you know much more open. I've always been quite you know sort of strong and controlled and managing things and you know I've had to ask for more help in this last year from anybody than I've ever done which has been, you know, sort of a learning curve, sort of unavoidable but sort of liberating at the same time. – P19, UK<br><br>I found Christmas very hard because my family were all around. My family, my three children are grown up. They're in their 20s and they were all back around and partners and children or whatever. I just… like I felt very like… I don't know. I felt very sad just for the way I was in that situation where I would be very much in the middle of that.– P9, Ireland |
| | Loss of friendships, relationships, social networks | I'm just glad I even can talk to you and you're my human contact for today. I don't have no other contacts for the rest of the day. That's why I just keep talking to you. It's like there's nobody else. I have nobody else for the rest of the day. – P39, Canada<br><br>With kind of friends or people who I thought were friends, it's stuff like 'oh but you were fine the other day' or kind of you know an expectation that you're doing something wrong. So 'have you been to the doctor's?' or… it's not kind of 'I do not believe you' but it's implied. – P32, UK |
| | Caregiving and social roles | If I had full custody of my kids, if I wasn't separated and sharing custody, there's no way that I'd have any level of improvement because my life just… as a single parent, if I was full time instead of 50 50, I wouldn't have the time to rest and recover from my caregiving duties. As it is, when my kids are in school, I spend my parenting weeks getting them out the door, resting and then doing my second shift when they get home. It takes all of my energy to do those things. It takes all of my energy to fulfil my parental obligations when they're around and then it takes the entire week when they're gone for me to recover and have to be in the place to repeat that cycle all over again. – P25, Canada |

**Supplemental File 2:** Dimensions of Disability in the Episodic Disability Framework in the Context of Long COVID - Supportive Quotes (n=40 participants)

| Dimension of Disability | Health challenges that comprise the dimensions | Supportive Quotes |
|---|---|---|
| | | It's affected my entire way of life. I'm now majorly housebound. I'm not going out and doing anything really. I can't even walk my baby around the block for fresh air just now. I have to… yeah, I have to wait for my partner to come in. He has to go and do that like he's doing just now. But yeah, just doing basic little things like that, it's a nightmare. Like walking up the stairs to bed, I can't walk upstairs with my baby just now because she's too heavy for me to carry as well as walking up the stairs. It's hard for me just to walk up the stairs on my own. So yes, it's huge. – P5, UK |
| | Work, employment, school | The thing with people with disabilities and what people don't get about people with disabilities is that yes, we're not working or some people for… like I guess I'm speaking for those who can't work or can't work very much right, is like some people might think it's like handouts and stuff like that. But honestly if we could work, we would work. We need some support and we need support that actually gives ability to afford everything that we need. Like again, I'm not looking for handouts but I do know that I need help. If I had capacity to work, then I would. So like I just don't want people to think that I have like some sort of entitlement thing just because I have a disability. – P38, Canada<br><br>It's not that we don't want to work. It's that we really can't and so we need things that are tailored to us and have that flex time and stuff like that… I can only do maybe a max of 10 hours a week if that. But through all that I'm still trying... it's taken a long time to find things but I'm still trying to find even that so that I can earn that little bit of extra income because I know even for me and part of it…feeling useful in society…I like to feel at least useful and helpful to people. So I haven't given up in trying to find at least a little bit of something but it's still I could never support myself. – P38, Canada |
| | Financial challenges (financial and housing insecurity and instability, student loans) | [Ontario Disability Support Program] ODSP is like six months to a year to get approved for… even if suddenly and miraculously I were approved for ODSP like tomorrow, which is not going to happen, I don't know how anybody lives on it. The idea with ODSP is that you get $1,169 [CAD] a month for rent, food, clothes and whatever you need. There's actually nowhere to live anywhere in Canada with that kind of money. And then they're like 'you can get a part time job' and I'm incapable of getting any kind of meaningful or you know… I can probably work an hour and a half every three days if I really tried hard and that job doesn't exist. So in that case you know I could get ODSP. I would still eventually become homeless and then they take away your shelter portion. So then … |

| Dimension of Disability | Health challenges that comprise the dimensions | Supportive Quotes |
|---|---|---|
| | | you have $600 a month to live on the street if you can imagine. So I don't have any financial uncertainty. What I have is a terrible fear of like homelessness. – P22, Canada<br><br>There's a pressure to maintain wellness, to keep well because I can't afford to not be well. – P1, UK<br><br>For most of this year it's been mainly just living off of savings and just cutting expenses here and there but having some financial support from family or you know my partner picking up a little bit here and there has been helpful. But yeah, I do stress about that and you know try to minimize the cost of things where I can. – P2, US |
| | Disruption to, and loss of retirement goals and planning | It's certainly not what I anticipated my retirement being. I had just completed… my retirement was going to include that mix of consulting. I had completed a yoga teacher training program. I was looking at different ski instructor programs, actually more assisted skiing, helping the disabled ski, being the guide for disabled skiers. So those are the things I was looking at. Those are gone you know, boom. – P15, US |
| **Uncertainty** | Uncertainty and worrying about the future; unpredictability of episodes, uncertainty about triggers of episodes, their sources, and treatments for Long COVID | So yeah, lots of uncertainty around what symptoms are going to happen you know. It's like I'll look down and be like that is very interesting, I don't know where that came from. Then obviously just like the general sort of you know the uncertainty of my life because of the fact that I can't work right. So yes, tons, all over the place. – P22, Canada<br><br>So there's a lot of uncertainty in terms of like from the health side of it of initially I was told in six months you'll be fine and we're not two years down the line and still not fine. – P30, UK<br><br>You see so many people recover and bounce back and back to their normal lives, friends of mine that got it at work. I just question sometimes why… and I'd love to know how I could have prevented from getting Long COVID or was it just one of these things they say are viral illnesses and it's not… I still would like to know, definitely. – P10, Ireland<br><br>I think not understanding why has the most psychological impact because you know you have the 'why me?' kind of mentality, no one's believing you and so you get angry and you get frustrated. If you understand at least the basics of why, you know the underlying mechanism of whatever is happening with you, then you can accept it and deal with it |

**Supplemental File 2:** Dimensions of Disability in the Episodic Disability Framework in the Context of Long COVID - Supportive Quotes (n=40 participants)

| Dimension of Disability | Health challenges that comprise the dimensions | Supportive Quotes |
|---|---|---|
| | | and then try and you know build your life to kind of pace yourself around it. But when you have no idea… – P1, UK<br><br>I'm still trying to figure out two years later what I'm capable of, although I am getting better or I feel like I am. But sometimes I feel like I plateau and how much is also getting better versus just better at pacing. – P24, US |
| | Worry about future health, ability to work, have a family, financial and housing security | I'm 48. I have lots of working time left in me. I'm in a job that suits me and yeah. I'm so uncertain about me being able to go back to that job at the level I was at. I was running a service. I couldn't even imagine myself in that situation again. Yeah, I don't know. There's so much uncertainty for me. I don't know where I would start. Everything is uncertain at the moment. – P9, Ireland<br><br>So there's kind of like am I going to go back into teaching. At the moment realistically no. You know, are we going to be able to afford a house, what am I going to do kind of job-wise and yeah. There's a lot of uncertainty, not just kind of financially and about my job but also you know am I going to be able to go on holiday again and you know enjoy things. So yeah, it's kind of having to change my mindset a lot especially in terms of what the future is going to look like because I don't really know. – P32, UK |
| | Implications for health and future life decisions (e.g. family, return to work) | But you know when I think about you know the future, especially my partner and I were thinking of having a baby, how you know would my body even be able to handle a pregnancy. I mean I can't handle a cold. You know like all these sorts of things obviously does affect my wellbeing. – P21, UK<br><br>I don't know if I'll be able to get back to work. Doing a very short walk like once in six or eight weeks is not going to be enough for me able to be fit enough to get back to do the job that I love. I know I'm on maternity leave at the moment and I will be until September. But feeling how I am at the moment, there's no way I could work, definitely not and I know that. Like I'm barely able to leave the house. So to actually go and help someone physically, it's a demanding job. – P5, UK<br><br>How will you know you're ready to return to work? [I've been told that] you have to be able to get through the basics of your own day to day life and be fully functional at it |

| Dimension of Disability | Health challenges that comprise the dimensions | Supportive Quotes |
|---|---|---|
| | | with capacity to spare before you can try and put your toe back into the working world. I haven't gotten there in the 14 months since my initial infection and it's really hard to know if I ever will. Maybe if treatments come along that can put in check those … crashes that I experience … Maybe we can find what the underlying cause is that keeps me sick and keeps me relapsing. Then maybe it will be conceivable. But as is I do not experience remittance for nearly long enough for that to be feasible…. It's just I'm not in a stable enough place for that to be a realistic goal. – P25, Canada |
| | Uncertainty experienced concurrently by health care providers, insurers and employers | As I'm getting the nearly 12 months now I am getting to get a bit worried like will I get back to where I was. I'd take 90%. If you offered it to me there, I'd take it. My doctor even said that to me. He said like there's no guarantee I'll get back to… I hope I can get back to 90%. I'd take that. – P10, Ireland<br><br>They opened the first long COVID clinic in DC where I am and I requested to go there because I had kind of given up on my doctor. Honestly, I sort of just like they didn't know what to do and I feel bad that I don't have the bandwidth for this. So my doctor was on board with… like she referred me there but then the insurance company wouldn't pay for it. – P26, US |

Note: This table does not include physical symptoms and impairments as these are reflected in the Table 1 of the main manuscript.
