## Supplementary material for "Conceptual framework of episodic disability in the context of Long COVID: Findings from a community-engaged international qualitative study": S3 - Contextual Factors of Disability - Supportive Quotes

**Supplemental File 3:** Contextual Factors (Extrinsic and Intrinsic) in the Episodic Disability Framework with Adults Living with Long COVID - Supportive Quotes (n=40 participants)

| **A - EXTRINSIC CONTEXTUAL FACTORS** | | |
|---|---|---|
| **Contextual Factor of Disability** | **Sub-categories of Contextual Factors** | **Supportive Quotes** |
| **Support (Practical, Emotional, Social) –** | Support from friends, family, partners (presence or absence) | Having people around me who reflect that pacing is good and will hold me accountable but also… it's such a hustle culture and that used to be my life and our society really glorifies like 'yeah you know sweat it out.' So being able to create this like environment for myself where these things but then aiming for are reflected back to me as positives just makes the whole thing so much easier. – P17, US<br><br>I know seeing family is a great thing to pick me up if I'm not feeling great. They always make me smile. I think some of it is that you can't be sad around a two year old and a five year old. Like just their keenness and their willingness to make me smile and make you happy no matter what and they'll draw me pictures and post them in my bedroom door if I'm sleeping. They are real little cuties. They're a good pick-me-up if I need or even just like looking at pictures or things like that is quite good if I need to… – P30, UK |
| | Support from Long COVID Community | If it wasn't for the online community, I don't even know where I'd be. I don't know because just being able to connect with people who believe you and are going through the same thing, who go the appointments and have the same bloody stupid things said to them, it helps. – P1, UK<br><br>…other people with Long COVID and some of them are health professionals. So that's been really helpful as well. One of them is a doctor who has Long COVID. So we've all been like kind of helping each other as well through it with our different skill sets. That's been really good. – P14, Canada |
| | Support from health and rehabilitation services and providers (presence or absence) | My physiotherapist as well, she's really good at bringing it all together and she speaks to everyone I think like in the hospital or the consultants. When I was on the respiratory ward they were like oh you're already having physiotherapy and they're like okay that's fine, do you mind if we speak to her. So that was really useful because they already knew each other, then they could build those links and sort of get the information shared that needs to be shared. – P30, UK<br><br>[My physiotherapist] would come back… and then we'd have another appointment, another session and he would say 'well I was thinking about you the other day.' So I |

| **A - EXTRINSIC CONTEXTUAL FACTORS** | | |
|---|---|---|
| **Contextual Factor of Disability** | **Sub-categories of Contextual Factors** | **Supportive Quotes** |
| | | was like this is amazing. He's actually looking to find solutions beyond just that one hour appointment that we have right. He's actually considering and looking and doing research beyond that one hour time slot that I have with him. I thought that's amazing and very comforting for me to think that this individual, this medical professional is curious as to what I have and wants to understand what it is if he can and to provide solutions based on the knowledge that we have gained together… throughout that time period he's always going back to finding out more information. – P16, Canada<br><br>My family doctor. She's very supportive but has no knowledge of post viral illness… very supportive in keeping me off work when I need it and listening to my needs. But not really coming up with any solutions besides just making sure that my blood tests are normal and making sure that I'm off work... almost all of the labour of finding adequate care has fallen on my shoulders. So I know the healthcare system. I know my way about it. But I've had to be extremely proactive in finding my own care. If I hadn't I would still be doing labs with my family doctor and that would be it. – P14, Canada |
| | Support from employers; insurers; human resource professionals; work colleagues (presence or absence) | Having a supportive boss has been huge. They were very understanding with sick days and being slower and they cut back hours. Like they really did a lot to try to help me stay in my job – P26, US<br><br>All my colleagues know that on any given day I may just say no, I can't do that and that's all they need to hear. They're also very understanding if I say I shouldn't be included in that project or not, I won't go to that meeting and I also say don't expect me to go to meetings alone… My colleagues are very good. They're not saying oh 'why can't you do that?', 'you're not pulling your weight'. No, that's been very good to have that understanding. – P15, US |
| | Support from program, policy, financial, income support; public health policies and mandates (presence or absence) | …there's also the huge financial strain. So my two years are fast approaching where I'm using my PRSI [Pay Related Social Insurance] stamps to get sick benefits. That will dry up and then I have nothing. So I already have no income and on you know social welfare benefits. They'll dry up. What then? So on top of everything else and the way I look at it is particularly the people who are in the first wave in March 2020, we've got a really hard blow because of the lack of testing, the false negatives and all that went with it and not being able to get a diagnosis with nobody knowing what it was, nobody |

| **A - EXTRINSIC CONTEXTUAL FACTORS** | | |
|---|---|---|
| **Contextual Factor of Disability** | **Sub-categories of Contextual Factors** | **Supportive Quotes** |
| | | even hearing of Long COVID. Like we were the beginnings, the first lot of people with Long COVID and to think that we're now still being discriminated against in terms of services. I'm scared to add up the bills I have in terms of what I paid to GPs and consultants because I've had to go private to all of them. I've had to go private to all of them. Otherwise I'd still be on indefinite waiting lists. I don't think that's fair that they're not recognizing what we have as a disability. – P36, Ireland<br><br>I'm drowning, quite frankly…every day I thought today is the day I'm going to get better, like this just can't last this long you know. People get better. You get sick and you get better. So I woke up every day with the full belief that any second now, like at any day that I woke up and I was okay a bit more mobility or I was in a better mood or I was like… I believed fully and completely that I was going to turn around any second. So you know I mean Long COVID isn't classified as a disability. You know even for to get EI you have to be available to work. I'm in a no-person's land you know. – P22, Canada |
| | Mobility aids, technology | Having access to digital technology has been so important to get everything from ordering my food, speaking to the GP, getting a taxi to the hospital. So that's key is having a phone or an internet connection. Anyone that doesn't, their quality of life, I don't know how they would be able to deal with Long COVID to be honest. – P3, UK<br><br>Up to December the previous year I was sort of making some improvement with my mobility especially. I'd been given a walker and I was you know able to go to the park and walk a bit without any major pain. So that was good. – P21, UK<br><br>I'm still having… I'm still suffering it because walking again is really tough. When I walk outside and… like I said when I walk I do use an aid. Well I've been using the wheelchair now as a prop so then I can sit down. – P21, UK |
| **Accessibility of environment and health services** | Accessibility of physical environment | Additional cognitive effort required to navigate environment - I would never have noticed, thought about or realised pre COVID. You just thought oh yeah there's a ramp, like wheelchair users can get in. It's the same with like dropped curbs. Everything is just a bit more of a challenge and you have to… it's kind of almost that extra cognitive effort to get anywhere because you have to be looking and aware of hold on a minute is that a good curb or a bad curb… – P30, UK |

| A - EXTRINSIC CONTEXTUAL FACTORS | | |
|---|---|---|
| **Contextual Factor of Disability** | **Sub-categories of Contextual Factors** | **Supportive Quotes** |
| | | I live in a house where it's not easily accessible. It's not on the flat…, the house is on different levels… I don't have a wheelchair either. But like in the kitchen now I would use a swivel chair which is something I use myself to get around the kitchen. – P37, Ireland<br><br>living in a place with stairs… it could be a challenge because I might go for weeks and weeks without being to go on the stairs. But equally, if I'm more alright like having an up period, then having the stairs means I can do some physical activity you know, especially because I don't go out as much. – P21, UK |
| | Physical accessibility of health services | Everyone's asking something of me thinking that that's going to be the trick that I'm going to get better when there's no definitive thing… everybody's experience is unique. Yes there are some things that are consistent that will help across people. But we all need different adaptations. We all need individualized support. – P38, Canada<br><br>I suppose, living in the country. Now like you have to drive to places. Trying to get to the hospital, I can't drive myself to the hospital anymore because there's no… services anywhere for Long COVID anywhere near where I live. So I have to travel to Dublin for the clinic I'm attending. It's not even a Long COVID clinic. It's under infectious diseases. But yeah, I have to get somebody to drive me there. – P35, Ireland |
| **Stigma and Epistemic Injustice** | Stigma and epistemic injustice enacted by family, friends, work colleagues, employers, insurers and health care providers;<br><br>Invisibility of Long COVID disability | I think it was about six months in is when I started to have cardiovascular issues, arrhythmias and tachycardia. It started about yeah month five or six. That turned out to be myocarditis. It took me over a year to get that diagnosed because nobody… when I first was sick and didn't get better, you know the thing was 'oh okay, get rest for weeks and this condition doesn't exist.' So it took me a very very long time to access any kind of healthcare. – P22, Canada<br><br>My initial attempts to get help were botched. I was told I was anxious. I was told that I was hiding a drug problem. I was told that it was menopause. So I couldn't get any kind of testing. I couldn't get anything done for a very long period of time. So I had no opportunity for early interventions. I was left alone… I know in terms of say healthcare or government agencies, two years is not a very long time. It's a really long time for me personally. Everyday is a nightmare.… my interactions with healthcare or any kind of |

| **A - EXTRINSIC CONTEXTUAL FACTORS** | | |
|---|---|---|
| **Contextual Factor of Disability** | **Sub-categories of Contextual Factors** | **Supportive Quotes** |
| | | support system have been brief and frustrating….I don't have a plan in place. I don't have a healthcare plan in place. - P35-Canada<br><br>I feel that it took me a little while to be believed. So initially I think it must have been about four weeks after I initially had COVID and I felt like something's not right. I know kind of in my gut something isn't right and I had kind of heard about long COVID. So I called the GP and spoke to one GP and she was like basically you're being dramatic, you're going to get over this. Then I tried again and had a similar thing and then I finally got through to a doctor who believed me and kind of referred me for different tests and stuff. So yeah, initially that was a bit of a challenge with just being believed I guess. – P32, UK<br><br>The most stressful part of the whole journey of Long COVID was fighting to be believed that I had COVID, that this was due to COVID and it wasn't pandemic stress related, that I had an actual physical problem… the sheer burden of trying to actually get seen and to get my symptoms being taken seriously caused so much emotional distress. There were so many times I just felt like giving up. The emotional or psychological trauma when you are not believed, when you know that something is wrong. You felt like you were going to die every night and then people are just dismissing you. – P1, UK<br><br>You look perfectly fine but you desperately need a seat. Like people aren't going to stand up for you just generally right. Like they'll just assume you'll be fine. So how do you go about that and what do you do when you're trying to get through like [name of store] to get medication and suddenly you're just too dizzy to stand – P39, Canada |

| **B - INTRINSIC CONTEXTUAL FACTORS** | | | |
|---|---|---|---|
| **Contextual Factor** | **Category of Contextual Factor** | **Sub-categories of Contextual Factors** | **Supportive Quotes** |
| **Living Strategies -** behaviors, attitudes and beliefs adopted to prevent, mitigate or deal with disability | **Maintaining control over health and life** | Pacing - anticipating, planning and preparing ahead; finding balance between activity and rest; navigating energy envelope; prioritizing, establishing structure and routine | Pacing is the [strategy]. It's the hardest one to stick to and you know it's been a work in progress... on the micro level… if you can't do it twice, don't do it once and not planning too much on any given day. How can I build in some scheduled breaks and also at a macro level of I'm used to being able to power through things? Now if I try to power through I get worse; how do I approach decisions like am I healthy enough to return to work with an eye to the need for pacing? – P17, US<br><br>All of these things made a big difference in the recovery and I felt a lot more in control of how to prevent symptoms coming. – P1, UK<br><br>There are people that I've avoided seeing because it's too much energy to try and explain [my disability to them]. – P19, UK<br><br>I need to plan my day and I always think … if I do something busy on a Monday, my Tuesdays are wiped off and then I'm thinking, do I need to do something Friday? Okay, well I need to probably have an easy day on Wednesday and Thursday because I know Friday is going to be quite draining or you know it depends on what I've got on." – P32 - UK |
| | | Lifestyle strategies (changes to diet, purposeful rest including sleep, stretching, mindfulness, taking up new life activities) | The satisfaction of realizing there's still something that I actually can do and I can create something and I can make something, so that's empowering. I find that hugely beneficial for me. – P36, Ireland<br><br>I can track my meals because I wasn't sure what foods were triggering me. I wasn't sure whether it was a certain number of steps or a certain temperature outside or what you know amount of activity was too much. So I had to get really granular about it… I went through a lot of trial and error with cutting out certain foods and cutting out a lot of foods, really limiting my activity to you know walks maybe at the ends of the day or even just tracking what chores I did that day whether it was just laundry or if I walked you know twice or whatever maybe. So I would track that on a day to day and then I |

| **B - INTRINSIC CONTEXTUAL FACTORS** | | | |
|---|---|---|---|
| **Contextual Factor** | **Category of Contextual Factor** | **Sub-categories of Contextual Factors** | **Supportive Quotes** |
| | | | would have you know a monthly calendar view of kind of the high points where I wouldn't need to track the meals necessarily but I might track the changes, I might track you know what supplements or treatments I was doing that day and what my overall score was for that day and anything that was different. So if I started something new or stopped something that I was doing I would log that. You know if I was trying to find some kind of pattern .. where I was getting better and better and better and then worse and worse and then better and then I could see what changes I made there and how I felt after each thing. – P2, US |
| | | Practical strategies (task modification, tracking health with wearables, journaling) | One of the things that has made it much more tangible for me is I have a Garmin VivoFit. I think a lot of us do. It calculates your body battery for the day, so like using heart rate variability and sleep and stress. It kind of calculates out of 100 how much energy you have and how much you used throughout the course of the day. Like for me at my sickest, like the spoons I had for today were like 7. Now like I can get into the 50s some days and that's pretty exciting. So just knowing like how much energy I have, what I want to spend it on. – P40, US<br><br>I'd say they're [triggers of disability] unpredictable. I've been trying to keep a diary each day. I've been trying to keep it hourly but I've not been 100% on top of that. … I'll feel tired but I still don't know what triggers the migraines. I don't know what makes the brain fog worse. … my heart rate can be completely normal but I'll feel like I can't breathe… so I'm trying to figure out what triggers it and I'm trying to take rests. … Sometimes it's just like the luck of the draw… I'm trying to figure out what's making me feel worse. – P32, UK<br><br>"One of the things that has made it much more tangible for me is I have a Garmin Vivofit. I think a lot of us do. It calculates your body battery for the day, so like using heart rate variability and sleep and stress. It kind of calculates out of 100 how much energy you have and how much you used |

| **B - INTRINSIC CONTEXTUAL FACTORS** | | | |
|---|---|---|---|
| **Contextual Factor** | **Category of Contextual Factor** | **Sub-categories of Contextual Factors** | **Supportive Quotes** |
| | | | throughout the course of the day. …. So just knowing how much energy I have, what I want to spend it on.” P67-US |
| | **Seeking social and practical support and knowledge** | Seeking out knowledge, health services and supports from others versus avoiding interactions with others | It sounds ridiculous but [asking for help is] actually a strategy that I would have avoided. You know I would inconvenience myself 10 times over rather than ask a friend to do something if it was going to be slightly difficult for them. Now I ask all the time. – P17, US<br><br>I’ve got family members that are like ‘you were really fit and active, like you can’t be still having symptoms’. I’ve got the point where I just don’t mention it anymore. I just think I don’t want to talk about it. I’ve only got a few close-knit friends that I will say I’m having a really bad day today. I’ve like had palpitations from 8am and it’s like yeah. It’s nice because they’re kind of… they’re going through the same thing. They’re like yeah I had that yesterday or… I’ve got a very good friend who’s a nurse and she’s absolutely brilliant. She used to be like yeah, I get it. – P4, UK<br><br>There are people that I’ve avoided seeing because it’s too much energy to try and explain [my disability to them]. – P19, UK |
| | **Attitudes, beliefs mindset and outlook** | Acceptance | At the beginning I felt more like I was you know definitely going to get better. It was probably around like, maybe around like the six month mark I had the first big realization that ‘oh shit I’m probably going to be dealing with this for the rest of the year’ you know. Then now at 15 months I mean that realization took a lot of different phases. At first it was like oh I’ll probably deal with this for a year. Then it was oh I’ll probably deal with this for another year. Now honestly where I’m at, I do have a lot of peace with it… I do have a lot of hope for a treatment just given the sheer amount of us that have gotten this you know post viral illness now. But when I look at the information that’s out there you know, other post viral illness patients, they are still sick 10, 20, 30, 40 years. So why would this virus be any different if I haven’t already gotten better and if I’m actually currently worse, you know? So it might sound depressing to you for me to say that but in all honesty it’s |

| **B - INTRINSIC CONTEXTUAL FACTORS** | | | |
|---|---|---|---|
| **Contextual Factor** | **Category of Contextual Factor** | **Sub-categories of Contextual Factors** | **Supportive Quotes** |
| | | | not depressing. It's an acceptance and it's allowed me to make decisions about my life to improve my life without living in this like liminal space where I'm waiting to 'get better.' So I feel like releasing that has given me a better quality of life. Like I said, I do still have a lot of hope for treatments. I just no longer have hope that I'm going to suddenly wake up and be at pre illness levels. – P34, US<br><br>I've learned that if I keep pushing, my body doesn't always catch up with what I want to do and you have to just let it… like just don't push too hard because it puts you back rather than if you take it a bit easier and build it slowly that you do make more progress. – P30, UK<br><br>After I stopped working I was able to rest a lot and also kind of came to the kind of trying to come to more of like an acceptance of like alright well these are the things that I can't do and these are the things that I can do and trying to work within my limitations. – P26, US |
| | | Adapting or adjusting mindset (shifting, adapting, adjusting and managing goals and expectations; patience; enhanced self-awareness; resiliency, taking every day as it comes) | Part of my journey I guess is patience - It's more like my body isn't catching up even though sometimes inside I feel like I'm normal again. – P23, US<br><br>Yeah, the biggest thing I say to people now is make sure you rest and pace because I've just learned the hard way too many times, that you can't trust your body and you can't push through it. You just really have to have patience. This condition has, you know it's taught me patience. – P29, Canada<br><br>Releasing that has given me a better quality of life… I do still have a lot of hope for treatments. I just no longer have hope that I'm going to suddenly wake up and be at pre illness levels. – P34, US<br><br>I learned in my mind to treat every day as a unique situation. If I wake up with little pain, then that is a blessing for me and I can then do the things that I need to do to be a good husband, a good father, a good employee and to |

| **B - INTRINSIC CONTEXTUAL FACTORS** | | | |
|---|---|---|---|
| **Contextual Factor** | **Category of Contextual Factor** | **Sub-categories of Contextual Factors** | **Supportive Quotes** |
| | | | make money so I can keep a roof over my head and feed my children" – P16, Canada |
| | | Hope and optimism; positive growth mindset (opportunity for growth) versus despair | There's this like type of pinecone that the seeds will only sprout if there's been a fire. You actually have to have a forest fire for like the seeds to come out. I feel like my life has like burnt to the ground … if I like look at it I'm a disabled single mom. Like it's not something I ever wanted in my 40s. But at the same time I'm becoming an advocate. Maybe I'm not going to get in the same… you know I had a lot of value in my job but I have lots of other talents. It's a way of like completely re-growing my life from like the start and there's nothing left. .. I get a chance to start over again which most people don't get, like they're kind of stuck in the rut of their life of what they chose. So I'm trying to see it as an opportunity to really spend the next half of my life with a lot of meaning… making meaningful choices and doing things that are meaningful. If I die I don't want to have any regrets. So how do I… Long COVID is an opportunity in some ways. I'm not saying I'm not sad but it's an opportunity to reinvent myself in some ways... – P14, Canada<br><br>I'm drowning, quite frankly… everyday I thought today is the day I'm going to get better, this just can't last this long you know. People get better. You get sick and you get better. So I woke up everyday with the full belief that any second now, like at any day that I woke up and I was okay a bit more mobility or I was in a better mood or I was like… I believed fully and completely that I was going to turn around any second… I'm in a no-person's land you know – P22, Canada. |
| | | Adopting new roles and purpose | It was the first thing that made it real because there are so many people that you know have it. The other thing is that I have found that I've been able to answer other peoples' questions and like bring in the ME and just say you know a b and c and say well I find this works for me that I found helped other people, more beneficial… I found helping other people when I could really helped at a time when I wasn't working. I think what I'm trying to say it |

| B - INTRINSIC CONTEXTUAL FACTORS | | | |
|---|---|---|---|
| **Contextual Factor** | **Category of Contextual Factor** | **Sub-categories of Contextual Factors** | **Supportive Quotes** |
| | | | helped with my self-esteem and it helped me like a purpose. I feel good about. In that way. – P11, Ireland<br><br>Feeling really passionate about the way that we think about and tell stories through data, through narratives but how do you around illness and the interplay between illness and mental health and physical health. I would love to explore ways in which I can continue to have a meaningful life that gives back to people in a way that I'm you know interested and qualified to do and expanding that capacity so that you know regardless of my physical state I can still do something good with my time. – P17, US |
| | **Diverting or Distracting** | | He said 'you've got to find the fun and once you find the fun, things will come from that.' So that's how creativity helped me because I had started drawing digitally. I've got an iPad. It meant I could… because I couldn't get out of bed. So I thought if I end up in the hospital at least I can look at things in bed. Then I started drawing one day and it was a distraction from everything. When I was able to express myself, I started keeping a diary and drawing. – P3, UK<br><br>I try not to think that I'm going to get back to pre-COVID days you know. – P17, US |
| **Personal Attributes** | Sex and gender identity | | I do wonder if there's a genetic element to a) catching it when you're exposed and b) then having long term issues. You know I think that peoples' bodies respond differently to it. There does seem to be a hormonal aspect here and women of all ages have reported that. – P19, UK<br><br>During the relapse, episodes around my menstruation time would be a massive trigger. So I nearly paced it to the… I'd know like four days beforehand, during the menstruation and then another week afterwards. So I would say 20 days of the month I'm nearly, I would say, housebound. – P6, Ireland |

| **B - INTRINSIC CONTEXTUAL FACTORS** | | | |
|---|---|---|---|
| **Contextual Factor** | **Category of Contextual Factor** | **Sub-categories of Contextual Factors** | **Supportive Quotes** |
| | Age | | I think people thought I would bounce back faster and there was just a lot of confusion around that as well because of the expectation that I should be able to just deal with it and that it wouldn't… so that has been quite… because the feedback constantly was about you're young, you're fit, you should be fine. – P3, UK |
|  | Ethnicity |  | I definitely think being white has been helpful to me in terms of being believed for my symptoms. I think being really educated and the job that I had … has also been helpful for being believed with my symptoms. – P26, US<br><br>Something that needs to be talked about as well is how much privilege I have as a white straight woman with middle class, with disability insurance, with a divorce from a rich man whose going to give me some money upon leaving me with… and I'm not going to starve to death. I have you know 65% of a good salary is still more than a lot of people earn…. it's not what I wanted in life but I'm not going to starve. My kid's fine. I'm not going to need social assistance. I'm educated. I have experience with the healthcare system. I think that with all this privilege that I have and I'm super conscious of and I find it really hard… – P14, Canada |
| | Pre-existing and other current health conditions | | I had IBS [Irritable bowel syndrome] pre COVID. So I go between the runs and constipation. First year of COVID it was completely the runs and second year is, the last six months, the bowels are not working. They're not functioning. It's not normal for me. This isn't my norm pre COVID. Yes I had sensitivities before to all meds typically. Even a B12 sublingual would put me in a full body rash. But now it's gone this high. Now I'm having anaphylactic shock and severe allergic reactions. My pharmacist knows me when I call him now, just my voice. That's pretty sad. – P13, Canada<br><br>When I look at the demographic data for long COVID, it seems like it is affecting more middle aged women on average or people with other underlying conditions or pre-existing conditions. – P2, US |

| **B - INTRINSIC CONTEXTUAL FACTORS** | | | |
|---|---|---|---|
| **Contextual Factor** | **Category of Contextual Factor** | **Sub-categories of Contextual Factors** | **Supportive Quotes** |
| | COVID characteristics (positive PCR test, length of time living with Long COVID) | | One of the things that helped me is that I had a positive test for COVID. So I was believed by the doctors – P14, Canada |
